# COVID-19 transmission in educational institutions August to December 2020 in Germany: a study of index cases and close contact cohorts

**DOI:** 10.1101/2021.02.04.21250670

**Authors:** Anja Schoeps, Dietmar Hoffmann, Claudia Tamm, Bianca Vollmer, Sabine Haag, Tina Kaffenberger, Kimberly Ferguson-Beiser, Berit Kohlhase-Griebel, Silke Basenach, Andrea Missal, Katja Höfling, Harald Michels, Anett Schall, Holger Kappes, Manfred Vogt, Klaus Jahn, Till Bärnighausen, Philipp Zanger

## Abstract

**Background:** The lack of precise estimates on transmission risk hampers rational decisions on closure of educational institutions during the COVID-19 pandemic.

**Methods:** Secondary attack rates (SARs) for schools and day-care centres were calculated using data from state-wide mandatory notification of SARS-CoV-2 index cases in educational institutions and information on routine contact tracing and PCR-testing.

**Findings:** From August to December 2020, every sixth of overall 784 independent index cases caused a transmission in educational institutions (risk 0·17, 95% CI 0·14–0·19). In a subgroup, monitoring of 14,594 institutional high-risk contacts (89% PCR-tested) of 441 index cases revealed 196 secondary cases (SAR 1·34%, 1·16–1·54). Transmission was more likely from teachers than from students/children (incidence risk ratio [IRR] 3·17, 1·79–5·59), and from index cases in day-care centres (IRR 3·23, 1·76–5·91) than from those in secondary schools. In 748 index cases, teachers caused four times more secondary cases than children (1·08 vs. 0·25 secondary cases per index, IRR 4·39, 2·67–7·21). This difference was mainly due to a large number of teacher-to-teacher transmissions in day-care centres (mean number of secondary cases 0.66) and a very low number of student/child-to-teacher transmissions in schools (mean number of secondary cases 0.004).

**Interpretation:** In educational institutions, the risk of infection for contacts to a confirmed COVID-19 case is one percent, but varies depending on type of institution and index case. Hygiene measures and vaccination targeting the day-care setting and teacher-to-teacher transmission are priorities in reducing the burden of infection and may promote educational justice during the pandemic.

**Funding:** No particular funding was received for this study.

**Research in context:** *Evidence before this study:* We searched PubMed on Jan 27, 2021, without any language restrictions for all articles in which the title or abstract contained the search terms “COVID 19” or “corona”, and “school”, “education*”, or “daycare”, and “transmission”, and “risk”, “attack rate”, or “SAR”, and screened 175 results for original research or reviews on COVID-19 transmission risk in the educational setting. Following a similar strategy, we also searched Google Scholar, SSRN, medRxiv, and the reference lists of identified literature. We found five cohort studies on transmission risk looking at overall 171 index cases and their 6,910 contact persons in Australian, Italian, Irish, Singaporean, and German schools and reporting attack rates between 0% and 3% percent. These five studies were conducted before October 2020 and thus looked at COVID-19 transmission risk in schools before the second wave in Europe. A number of modelling studies from the first wave of COVID-19 provide inconclusive guidance to policy makers. While two publications, one from several countries and one from Switzerland, concluded that school closures contributed markedly to the reduction of SARS-CoV-2 transmission and individual mobility, two other studies, one using cross-country data and one from Japan rated school closures among the least effective measures to reduce COVID-19 incidence rates.

*Added value of this study:* Based on a large data set that emerged from the current public health practice in Germany, which incorporates routine PCR-testing during active follow-up of asymptomatic high-risk contacts to index cases, this study provides a precise estimate of the true underlying SARS-CoV-2 transmission risk in schools and day-care centres. Its analysis also allows for a meaningful examination of differences in the risk of transmission with respect to the characteristics of the index case. We found that the individual risk of acquiring SARS-CoV-2 among high-risk contacts in the educational setting is 1·3%, but that this risk rises to 3·2% when the index case is a teacher and to 2·5% when the index case occurs in a day-care centre. Furthermore, we could show that, on average, teacher index cases produced about four times as many secondary cases as student/child index cases. Despite the relatively small proportion of teachers among index cases (20%), our study of transmission pathways revealed that the majority of all secondary cases (54%), and the overwhelming majority of secondary cases in teachers (78%) were caused by teacher index cases. Of note, most cases of teacher-to-teacher transmission (85%) occurred in day-care centres.

*Implications of all the available evidence:* In this setting, where preventative measures are in place and COVID-19 incidence rates were rising sharply in the population, we found a low and stable transmission risk in educational institutions over time, which provides evidence for the effectiveness of current preventative measures to control the spread of COVID-19 in schools. The identification of a substantial teacher-to-teacher transmission risk in day-care, but a clearly mitigated child/student–to-teacher transmission risk in schools, indicates the need to shift the focus to hygiene among day-care teachers, including infection prevention during staff-meetings and in break rooms. These findings also strongly support the re-prioritization of vaccination against SARS-CoV-2 to educational staff in day-care.

## Introduction

The 2020 COVID-19 pandemic urges government leaders to define priorities when implementing anti-epidemic measures in public domains. This task requires a profound scientific basis on the beneficial and hazardous effects of these restrictions on social, economic, and health outcomes. Since the start of the colder season in the Northern Hemisphere, the number of infections with COVID-19 has started to rise again, reaching more than half a million newly reported cases globally per day in December 2020.^1^ These alarmingly high numbers have led decision makers worldwide to impose partial or complete lockdowns. In the majority of countries, these measures include the full or partial closure of educational institutions.^2^

The implications of school closures are multifaceted and are considered to affect children from most deprived families the hardest^3^, with limited access to computers and the internet as one out of several mechanisms mediating this disadvantage. Even under the best conditions, home schooling with the parents, online teaching, TV education, or a combination thereof, are inferior learning environments as opposed to the classroom context. Besides, as schools also fulfil the task of child care, school closures not only affect the children’s learning and mental health^4^, but also the parents’ ability to pursue their work obligations. Similar reasoning applies to day-care facilities. Furthermore, the transfer of child care duties to grandparents increases the risk of COVID-19 for an older population group, which will be associated with a higher number of severe disease and eventually COVID-19-related deaths.^5^ This well-established perception on the educational shortcomings of school closures is contrasted by incomplete knowledge on COVID-19 transmission during onsite education. Accordingly, a recent review on SARS-CoV-2 setting-specific transmission rates concluded that there is “limited data to explore transmission patterns in […] schools […], highlighting the need for further research in such settings.” ^6^ (see research in context for more detail).

Against this background, this study provides information on SARS-CoV-2 transmission risk and patterns in schools and day-care centres during a period with exponentially increasing COVID-19 population incidence. This research makes use of data that emerged between August and December 2020 from containment measures around index cases in educational institutions that were mandated by District Public Health Authorities (DPHAs) in Rhineland-Palatinate, Germany.

## Methods

### Source population

The presented data was collected in Rhineland-Palatinate, one of the 16 Federal States of Germany with an overall population of about 4.1 million, 1492 schools, 406,607 school-children, and 144,245 children below 6 years of age in day-care.^7,8^ We report observations from the re-opening of educational institutions after the summer break, on August 17^th^, to their closure for a hard lock-down, on December 16^th^, 2020. During this period, publicly recommended hygiene measures in secondary schools (i.e. beginning approximately with age 10 years) in Rhineland Palatinate included (i) physical distancing (> 1.5 meters), (ii) cross- or pulse-ventilation of class-rooms before and after class, and then every 20 minutes for 5 minutes during class,^9^ (iii) face masks in school-buildings and “on campus”, but not in the class-room, (iv) increased frequency of surface cleaning, and (v) structural support of individual hygiene (hand, cough etiquette).^10,11^ On November 2^nd^, 2020, this concept was modified by additionally recommending face masks inside the classroom.^11^ Comparable recommendations existed for primary schools and day-care centres, but exempting the children from physical distancing (i) and wearing of face masks (iii).^6^

SARS-CoV-2 and COVID-19 are notifiable according to the German Infectious Diseases Protection Act (Infektionsschutzgesetz, IfSG), a Federal Law implemented by each Federal State’s own jurisdiction. Based hereon, physicians and laboratories have to notify COVID-19 cases to the responsible DPHA alongside with contextual information. This includes information on whether the person is a child or teacher in a school or day-care centre (§33 IfSG). From the DPHAs, information on newly identified COVID-19 cases is forwarded to one of the 16 Federal State Infectious Diseases Surveillance Centers (Landesmeldestellen), within 24 hours, and from there to the national surveillance centre at Robert Koch-Institute (RKI) in Berlin.

### Origin of index cases and contact persons

Each of the 24 DPHAs in Rhineland-Palatinate is responsible for all investigations around notifiable diseases, and represent populations between 61,000 and 430,000 individuals per district. Following the identification of a COVID-19 case, qualified personnel at the competent DPHA interviews the index case, traces contacts, and initiates an active follow-up of those in category-I for fourteen days following the last contact with the index case. A category-I contact is defined as a person who either stayed face-to-face (<1.5 meters) with a COVID-19-case for 15 minutes or longer, or in the same room (i.e. irrespective of distance) for 30 minutes or longer.^12^ In the context of schools and day-care centres, the Robert Koch-Institute recommends that DPHAs classify all members of a class or group as category-I contact persons in crowded or unclear situations, or when resources at the DPHA do not allow for an individual risk assessment.^10^ Furthermore, according to current RKI guidelines, free PCR-testing is offered by the DPHAs to all category-I contact persons, irrespective of their symptom status.^12^ Depending on the DPHAs organizational structure, the testing is organized by the DPHA personnel or by external structures, such as community testing centres. In the latter case, only SARS-CoV-2 positive PCR-tests of secondary cases are notified to the DPHA, with the result that negative test results are only available to the DPHAs who organize testing themselves. This explains why DPHAs with external testing have missing information on the total number of tests done in contact persons of a given index cases, even when such testing was routinely offered to all contact persons as a standard procedure for all index cases included in this study.

### Study setting and definitions

With start of the study, a 2-page questionnaire was distributed to all DPHAs, alongside with detailed instructions on inclusion and exclusion criteria for index cases and a link to upload completed questionnaires (ec.europa.eu/eusurvey). For inclusion into this study, an index case was defined as an individual that (i) tested positive for SARS-CoV-2-RNA from respiratory material, (ii) was notified as working in or attending an educational institution according to §33 IfSG, and (iii) had worked in or attended the institution for at least one day during the infectious period. The infectious period was defined as follows: (i) for symptomatic index cases as time from 2 days before to 10 days after onset of symptoms; (ii) for asymptomatic cases with unknown origin from 2 days before to 10 days after the date of taking the diagnostic swab; and (iii) for asymptomatic cases with known contact to a primary case from 3 days to 15 days after exposure.

A secondary case was defined as an individual that was identified (i) as a contact person to an index case by the competent DPHA and (ii) tested positive for SARS-CoV-2-RNA during the quarantine associated with that index case. Contact persons and secondary cases not attending the educational institution, e.g. persons living in the same household, were not to be reported in this questionnaire. Beginning with August 17^th^, 2020, we asked DPHAs to file one questionnaire for each eligible index case about 2 weeks after its identification, when information on all potential secondary cases would be available, i.e. after completion of quarantine of the contact persons. Afterwards, DPHAs received requests for contribution to SARS-S on a weekly basis, alongside interim analysis of the surveillance.

### Statistical analysis

Assuming equal exposure time in all contact persons of a given index case, we calculated secondary attack rates (SAR, i.e. individual level risk of transmission) as the proportion of secondary cases among contact persons of a given index case, together with corresponding binomial 95% confidence intervals (95% CIs).^13^ Associations between transmission risk on the individual level and a number of the index case’s characteristics were analysed using negative binomial regression, providing estimates of the crude incidence risk ratio (IRR) comparing the SAR in exposed and unexposed together with its 95% CI and a p-value testing H0: “both SARs are equal” (IRR=1.0). For sensitivity, we repeated these analyses based on the number of PCR-tested contacts only.

The risk of causing a SARS-CoV-2 cluster was calculated by dividing the number of index cases with at least one secondary case by the total number of index cases. To this end, we used binomial regression models estimating risk ratios (RRs) with 95% CIs and p-values testing H0: RR=1.0. Since this part of the analysis did not rely on an individual person denominator, it also included data from index cases with missing information on total number of contact persons and/or number of PCR tests done. Finally, we used negative binomial regression to estimate IRRs, their 95% CIs, and corresponding p-values for comparing the proportions of secondary cases caused by students/children and teachers, respectively. All analyses were conducted in Stata SE version 16.1 and SAS version 9.4.^14,15^

### Ethical statement

The collection, analysis and communication of the presented data take place in response to the global COVID-19 pandemic and are mandated by the German Infectious Diseases Protection Act. Ethical approval was waived by the competent ethics committee, Federal State Medical Council (Landesärztekammer) in Rhineland-Palatinate, Mainz, Germany (application no. 2021-15634-r).

## Results

### Source population

The course of the COVID-19 pandemic in Rhineland-Palatinate was comparable to all of Germany and was characterized by an exponential increase from the end of September (calendar week 39) until the end of October (cw43), a further growth at a lower rate (cw44–45), followed by a fluctuation on a high level of about 5000 to 7000 new cases per week (cw46–52), which equals a 7-day incidence rate of 120 to 170 per 100,000 (figure 1). Sixteen percent of the 74,733 COVID-19 cases notified in Rhineland-Palatinate in 2020 were younger than 20 years, which approximates the population proportion of 18.3% in this age group. Among these, 1,954 notifications contained contextual information on educational institutions (1,298 students/children and 684 teachers), 84% of which were notified during the study period.

**Figure 1:**
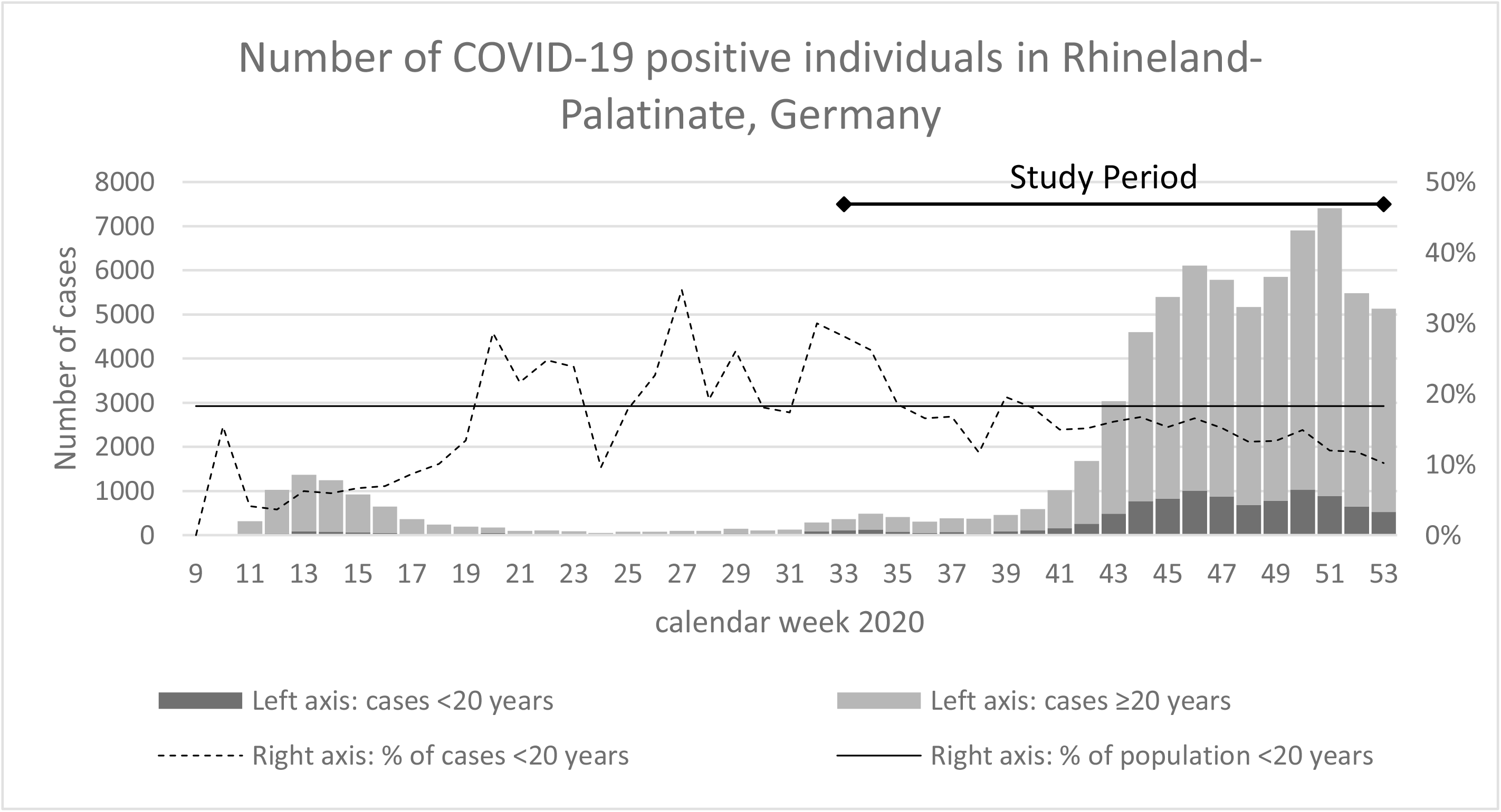
Age-stratified course of the COVID-19 pandemic in Rhineland-Palatinate, Germany, 2020. Figure displays number (%) of SARS-CoV-2 cases notified in the Federal State of Rhineland-Palatinate by calendar week, overall, and among subjects ≤ 20 years of age.

### Secondary attack rates (SAR)

Overall, DPHAs provided additional information for a total of 784 independent notified index cases attending an educational institution prior to diagnosis of a SARS-CoV-2 infection (591 students/children, 157 teachers, 36 unknown role). Full information on PCR-testing was available for 14,591 contact persons to 441 index cases (median 25 contacts per index case, IQR 17-40). Among these, the DPHAs identified 81 clusters with 196 PCR-positive secondary cases (SAR 1.34%, 95% CI 1.16%–1.54%) (table 1, left). Repeating the analysis based on only the 13,005 PCR-tested contacts (PCR-coverage 89%), gave an overall SAR of 1.51 (95% CI 1.30–1.73). The majority of contacts (71%) were tested between seven and ten days after their last contact with the index case (supplementary figure 1).

**Table 1:** Risk of COVID-19 transmission in educational settings, by characteristic of the index case, Rhineland-Palatinate, Germany, 2020

| Characteristic of index case | Risk of transmission on individual level (SAR), N=441 | | | | | | | Risk of transmission on cluster level ($\geq 1$ secondary case), N=784 | | | | | |
| --- | --- | --- | --- | --- | --- | --- | --- | --- | --- | --- | --- | --- | --- |
|  | Index cases<sup>a</sup> | Secondary cases (clusters) | No. of contacts | % Contacts with PCR | SAR (%) [95% CI] | Crude IRR [95% CI] | P-value | Index cases<sup>b</sup> | Clusters (secondary cases) | Mean cluster size | Risk [95% CI] | Crude RR [95% CI] | P-value |
| **Role** |  |  |  |  |  |  |  |  |  |  |  |  |  |
| Student/child | 346 | 99 (53) | 10716 | 87.47 | 0.92 [0.75–1.12] | 1.00 | ref | 591 | 76 (145) | 1.9 | 0.13 [0.10–0.16] | 1.00 | ref |
| Teachers | 75 | 91 (25) | 2858 | 92.93 | 3.18 [2.57–3.90] | 3.17 [1.79–5.59] | <0.001 | 157 | 48 (169) | 3.5 | 0.31 [0.23–0.38] | 2.38 [1.73–3.26] | <0.001 |
| unknown | 20 | 6 (3) | 1017 | 95.97 | 0.59 [0.22–1.28] | 0.77 [0.23–2.63] | 0.68 | 36 | 6 (15) | 2.5 | 0.17 [0.06–0.33] | 1.30 [0.61–2.77] | 0.50 |
| **Symptom status** |  |  |  |  |  |  |  |  |  |  |  |  |  |
| Symptomatic | 300 | 166 (64) | 10566 | 88.11 | 1.57 [1.34–1.83] | 1.00 | ref | 550 | 100 (262) | 2.6 | 0.18 [0.15–0.22] | 1.00 | ref |
| Asymptomatic | 127 | 21 (13) | 3523 | 91.26 | 0.60 [0.37–0.91] | 0.47 [0.25–0.89] | 0.02 | 203 | 24 (55) | 2.3 | 0.12 [0.08–0.17] | 0.65 [0.43–0.99] | 0.04 |
| unknown | 14 | 9 (4) | 502 | 95.62 | 1.79 [0.82–3.38] | 1.34 [0.37–4.85] | 0.65 | 31 | 6 (12) | 2.0 | 0.19 [0.07–0.37] | 1.06 [0.51–2.23] | 0.87 |
| **Type of institution<sup>c</sup>** |  |  |  |  |  |  |  |  |  |  |  |  |  |
| Day-care (0-6 years) | 99 | 110 (32) | 4392 | 90.64 | 2.50 [2.06–3.01] | 3.23 [1.76–5.91] | <0.001 | 205 | 61 (203) | 3.3 | 0.30 [0.24–0.37] | 2.78 [1.88–4.10] | <0.001 |
| Primary Schools | 88 | 27 (15) | 2389 | 85.64 | 1.13 [0.75–1.64] | 1.62 [0.80–3.31] | 0.18 | 157 | 21 (40) | 1.9 | 0.13 [0.08–0.20] | 1.25 [0.75–2.09] | 0.40 |
| Secondary Schools | 173 | 41 (25) | 5970 | 90.08 | 0.69 [0.49–0.93] | 1.00 | ref | 299 | 32 (50) | 1.6 | 0.11 [0.07–0.15] | 1.00 | ref |
| Vocational Schools | 52 | 5 (4) | 1181 | 84.17 | 0.42 [0.14–0.99] | 0.65 [0.21–1.99] | 0.45 | 70 | 8 (14) | 1.8 | 0.11 [0.05–0.21] | 1.07 [0.51–2.22] | 0.18 |
| Other/unknown | 29 | 13 (5) | 659 | 91.96 | 1.97 [1.05–3.35] | 2.99 [1.12–7.98] | 0.03 | 53 | 8 (22) | 2.8 | 0.15 [0.07–0.28] | 1.41 [0.69–2.89] | 0.35 |
| **Age** |  |  |  |  |  |  |  |  |  |  |  |  |  |
| 0-5 years | 42 | 31 (11) | 1828 | 90.43 | 1.70 [1.16–2.40] | 1.29 [0.55–3.03] | 0.55 | 77 | 18 (45) | 2.5 | 0.23 [0.14–0.34] | 2.00 [1.14–3.53] | 0.02 |
| 6-10 years | 89 | 15 (12) | 2410 | 82.53 | 0.62 [0.35–1.02] | 0.72 [0.32–1.63] | 0.43 | 155 | 18 (31) | 1.7 | 0.12 [0.07–0.18] | 1.00 [0.56–1.79] | 0.99 |
| 11-15 years | 113 | 35 (20) | 3358 | 89.93 | 1.04 [0.73–1.45] | 1.00 | ref | 189 | 22 (38) | 1.7 | 0.12 [0.07–0.17] | 1.00 | ref |
| 16-20 years | 90 | 17 (9) | 2884 | 88.04 | 0.59 [0.34–0.94] | 0.56 [0.25–1.24] | 0.15 | 157 | 16 (37) | 2.3 | 0.10 [0.06–0.16] | 0.88 [0.48–1.61] | 0.67 |
| 21-34 years | 42 | 30 (13) | 1321 | 88.19 | 2.27 [1.54–3.23] | 1.97 [0.84–4.64] | 0.12 | 80 | 23 (50) | 2.2 | 0.29 [0.19–0.40] | 2.47 [1.46–4.17] | 0.001 |
| 35 years and older | 45 | 62 (13) | 1773 | 93.80 | 3.50 [2.69–4.46] | 2.80 [1.27–6.20] | 0.01 | 90 | 27 (113) | 4.2 | 0.30 [0.21–0.41] | 2.58 [1.56–4.27] | <0.001 |
| unknown | 20 | 6 (3) | 1017 | 95.97 | 0.59 [0.22–1.28] | 0.65 [0.18–2.35] | 0.51 | 36 | 6 (15) | 2.5 | 0.17 [0.06–0.33] | 1.43 [0.62–3.28] | 0.40 |
| **Type of groups** |  |  |  |  |  |  |  |  |  |  |  |  |  |
| Stable Groups | 313 | 80 (52) | 8650 | 88.15 | 0.92 [0.73–1.15] | 1.00 | ref | 536 | 75 (123) | 1.6 | 0.14 [0.11–0.17] | 1.00 | ref |
| Dynamic Groups | 111 | 91 (25) | 5430 | 90.90 | 1.68 [1.35–2.05] | 1.45 [0.84–2.50] | 0.18 | 199 | 44 (168) | 3.8 | 0.22 [0.17–0.29] | 1.58 [1.13–2.21] | 0.007 |
| unknown | 17 | 25 (4) | 511 | 86.89 | 4.89 [3.19–7.14] | 3.21 [1.07–9.59] | 0.04 | 49 | 11 (38) | 3.5 | 0.22 [0.12–0.37] | 1.60 [0.92–2.81] | 0.10 |
| **Sex** |  |  |  |  |  |  |  |  |  |  |  |  |  |
| Female | 194 | 66 (33) | 5808 | 88.86 | 1.14 [0.88–1.44] | 1.00 | ref | 332 | 43 (89) | 2.1 | 0.13 [0.10–0.17] | 1.00 | ref |
| Male | 241 | 128 (47) | 8555 | 89.32 | 1.50 [1.25–1.78] | 0.78 [0.46–1.31] | 0.35 | 441 | 86 (238) | 2.8 | 0.20 [0.16–0.24] | 0.66 [0.47–0.93] | 0.02 |
| unknown | 6 | 2 (1) | 228 | 89.04 | 0.88 [0.11–3.13] | 0.64 [0.07–5.67] | 0.69 | 11 | 1 (2) | 2.0 | 0.09 [0.0–0.41] | 0.47 [0.07–3.05] | 0.43 |
| **Month of infection** |  |  |  |  |  |  |  |  |  |  |  |  |  |
| August | 33 | 6 (5) | 909 | 97.36 | 0.66 [0.24–1.43] | 1.00 | ref | 33 | 5 (6) | 1.2 | 0.15 [0.05–0.32] | 1.00 | ref |
| September | 78 | 9 (9) | 2548 | 97.84 | 0.35 [0.16–0.67] | 0.45 [0.12–1.78] | 0.26 | 78 | 9 (9) | 1.0 | 0.12 [0.05–0.21] | 0.76 [0.28–2.10] | 0.60 |
| October | 95 | 84 (23) | 3986 | 93.15 | 2.11 [1.68–2.60] | 1.92 [0.58–6.42] | 0.29 | 149 | 24 (85) | 3.5 | 0.16 [0.11–0.23] | 1.06 [0.44–2.58] | 0.89 |
| November | 151 | 74 (30) | 4492 | 85.28 | 1.65 [1.30–2.06] | 1.77 [0.55–5.75] | 0.34 | 349 | 56 (150) | 2.7 | 0.16 [0.12–0.20] | 1.06 [0.46–2.46] | 0.89 |
| December | 84 | 23 (14) | 2656 | 78.43 | 0.87 [0.55–1.30] | 1.11 [0.32–3.91] | 0.87 | 175 | 36 (79) | 2.2 | 0.21 [0.15–0.27] | 1.36 [0.58–3.20] | 0.49 |
| **Total** | **441** | **196 (81)** | **14591** | **89.13** | **1.34 [1.16–1.54]** | **n.a.** | **n.a.** | **784** | **130 (329)** | **2.5** | **0.17 [0.14–0.19]** | **n.a.** | **n.a.** |

Left side of table displays secondary attack rates (SARs), defined as the proportion of secondary cases in all contact persons. Incidence risk ratios (IRRs) and corresponding confidence intervals (CIs) and p-values from negative binomial regression compare the SARs between different groups. The right side of the table displays the cluster risk, i.e. the risk of causing at least one secondary infection, and associated risk ratios (RRs) from binomial regression for the comparison between groups. <sup>a</sup>Index cases with information on number of contact persons and number of PCR tests, <sup>b</sup>complete study population, <sup>c</sup>includes teachers

The SARs varied by characteristic of the index case: role (teacher > student/child, IRR=3.17, p<0.001), symptom status at time of diagnosis (pre-/asymptomatic < symptomatic, IRR=0.47, p=0.02), type of institution (day-care centres > secondary schools, IRR=3.23, p<0.001), and by age (older than 35 years > age between 6 and 21 years) (table 1, left).

### Cluster level risk and cluster size

To calculate the risk of causing a cluster (i.e. the risk of transmission to at least one secondary case), we included additional 343 index cases with complete information on secondary cases, but missing information on total number of contact persons tested. In the combined dataset of overall 784 index cases, there were 130 clusters reported via the SARS-Surveillance (cluster risk 0.17, 95% CI 0.14–0.19). We observed variation in risk of causing a cluster by characteristic of the index case (i.e. role, symptom status, type of institutions, and age) that was very similar in direction and magnitude to the comparisons based on the SARs. Index cases in dynamic groups were more likely to cause SARS-CoV-2 clusters than index cases in stable groups.

### Transmission patterns by role of index case

Teacher index cases caused on average more secondary cases (169/157, risk=1.08) than students/children (145/591, risk=0.25; IRR 4.39, p<0.001) (table 2). Assessing transmission patterns by role of index and secondary cases, we found that the average number of student/child-to-teacher transmission was 0.04 (corresponding to about one teacher secondary case in 25 student/child index cases) compared to 0.56 for teacher-to-teacher transmission (one teacher secondary case in 2 teacher index cases, IRR 13.25, p<0.0001). A similar comparison looking at secondary cases in children/students found a similar but less pronounced difference towards a more likely teacher-to-student/child transmission (81/157, risk=0.52) compared to transmission from students/children to peers (120/591, risk=0.20, IRR 1.54, p<0.001). Looking at role-specific transmission-patterns while stratifying by type of institution identified a comparably high transmission risk from teacher indexes in day-care (on average 1.26 secondary cases per index case) to both, teachers (0.66) and children (0.59). In schools, increased transmission risk from teacher indexes (0.50) was only reproducible with regard to student (0.44), but not to teacher secondary cases (0.06). Transmission from child indexes in day-care (0.66) was about equally likely to peers (0.38) than to teachers (0.28). Student indexes in schools (0.17), by contrast, hardly ever transmitted to teachers (0.004) (table 2). Figure 2 displays transmission in absolute numbers, attributable to the proportion of the index cases’ roles and institutions in the study sample.

**Table 2.** COVID-19 transmission between students/children and teachers, Rhineland-Palatinate, Germany, 2020 (N=748)^a^

| Transmission from |  | to all types of secondary cases (n=314)<sup>a</sup> |  |  |  | to student/child (n=201) |  |  |  | to teachers (n=113) |  |  |  |
| --- | --- | --- | --- | --- | --- | --- | --- | --- | --- | --- | --- | --- | --- |
|  |  | N cases (clusters) | Risk<sup>b</sup> | IRR [95% CI]<sup>c</sup> | P-value<sup>c</sup> | N cases (clusters) | Risk<sup>b</sup> | IRR [95% CI]<sup>c</sup> | P-value<sup>c</sup> | N cases (clusters) | Risk<sup>b</sup> | IRR [95% CI]<sup>c</sup> | P-value<sup>c</sup> |
| **All institutions** | student/child, n=591 | 145 (76) | 0.25 | 1.00 |  | 120 (70) | 0.20 | 1.00 |  | 25 (16) | 0.04 | 1.00 |  |
|  | teachers, n=157 | 169 (48) | 1.08 | 4.39 [2.67–7.21] | <0.001 | 81 (37) | 0.52 | 2.54 [1.52–4.23] | <0.001 | 88 (32) | 0.56 | 13.25 [6.59–26.67] | <0.001 |
| **Day-care** | student/child, n=79<sup>d</sup> | 52 (20) | 0.66 | 1.00 |  | 30<sup>d</sup> (15) | 0.38 | 1.00 |  | 22<sup>d</sup> (13) | 0.28 | 1.00 |  |
|  | teachers, n=113<sup>d</sup> | 142 (38) | 1.26 | 1.91 [0.95–3.84] | 0.07 | 67<sup>d</sup> (32) | 0.59 | 1.56 [0.78–3.13] | 0.21 | 75<sup>d</sup> (26) | 0.66 | 2.38 [1.04–5.49] | 0.04 |
| **Schools** | student/child, n=474<sup>d</sup> | 82 (52) | 0.17 | 1.00 |  | 80<sup>d</sup> (51) | 0.17 | 1.00 |  | 2<sup>d</sup> (2) | 0.004 | 1.00 |  |
|  | teachers, n=32<sup>d</sup> | 16 (6) | 0.50 | 2.89 [0.99–8.44] | 0.05 | 14<sup>d</sup> (5) | 0.44 | 2.59 [0.86–7.83] | 0.09 | 2<sup>d</sup> (2) | 0.06 | 14.81 [2.09–105.15] | 0.007 |

Table displays the risk of transmission calculated as mean number of secondary cases per index case, conditional on the role of the index case (i.e. student/child vs. teacher), and stratified by type of institution (day-care centre, school). For instance, independent of institution, the mean number of secondary cases in teachers are 0.04 and 0.56 for student/child and teacher index cases, respectively, corresponding to an IRR of 13.3; the same comparison for day-care centres results in an IRR of 2.4. <sup>a</sup>36 index cases with missing age/role information, <sup>b</sup>mean number of new cases per index case, <sup>c</sup>Incidence Risk Ratios (IRRs) and corresponding confidence intervals (CIs) and p-values from negative binomial regression, <sup>d</sup>number of subjects do not add up to total due to exclusion of cases that attended institutions other than schools and day-care centres (e.g. special needs schools).

**Figure 2:**
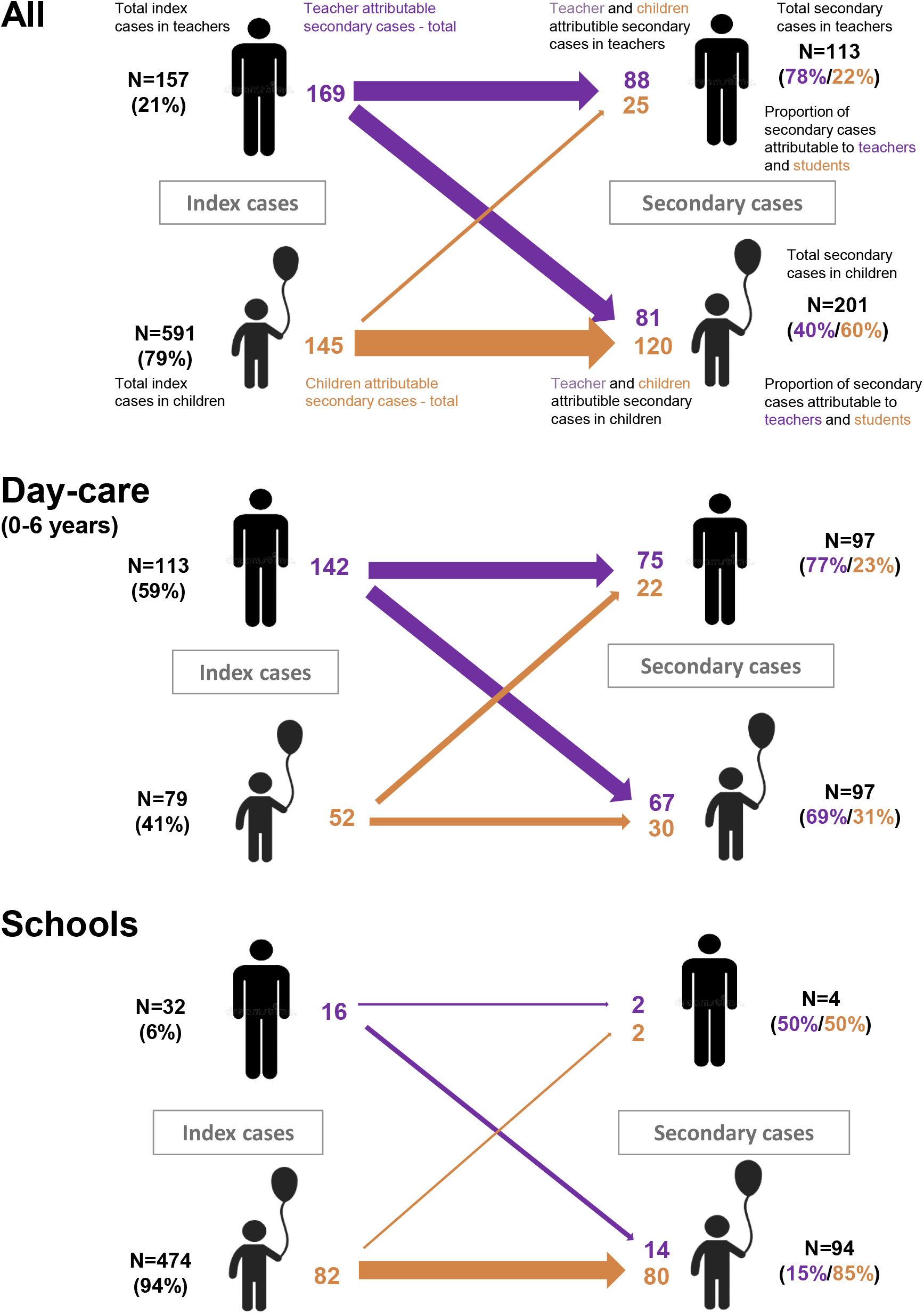
Transmission pathways in educational institutions, as total number of secondary cases, from teacher- and child/student-index cases to teacher and child/student secondary cases, by type of institution. Width of arrows is proportional to absolute number of secondary cases per index

### Index case, cluster size, and cluster-composition

The 329 secondary cases reported in this study occurred in 130 clusters, while the majority, 654 of overall 784 indexes (83%), led to zero secondary cases. In those 130 cases, where transmission occurred, the average cluster size was 2.5 secondary cases (supplementary figure 2). There were nine clusters reported with seven or more secondary cases (table 3), of which seven (78%) were caused by a teacher index. Seven of the large outbreaks were in day-care centres for young children, where the index cases had on average 78 category-I contacts. All nine outbreaks occurred in settings, where the index cases had a large number of category-I contacts (between 37 and 166 contacts), as opposed to an average of 33 (median 25) contacts per index case in the overall study. Larger outbreaks were more often caused by teachers (mean cluster size 3.5 vs 1.9) explaining why “black dots” prevail when moving from the lower left to the upper right corner of the grid in figure 3. Outbreaks involving several teachers follow commonly on an index case in teachers and rarely on an index in children/students, explaining why there are only a few white dots with proximity to the y-axis and clearly more white dots closer to the x-axis.

**Table 3:** Specific information on the nine largest clusters in our study (size ≥ 7)

|  | Month of Symptom Onset<sup>a</sup> | Role of Index | Age group of Index | Type of Institution | Number of high risk contacts<sup>b</sup> |  | Number of Secondary Cases |  |  |
| --- | --- | --- | --- | --- | --- | --- | --- | --- | --- |
|  |  |  |  |  | Total | % PCR-tested | Total | Children | Teachers |
| Cluster 1 | December | Teacher | 41-45 | Day-care $\leq 6$ years | 43 | Unknown | 8 | 3 | 5 |
| Cluster 2 | October | Child | 0-5 | Day-care $\leq 6$ years | 150 | 70% | 9 | 7 | 2 |
| Cluster 3 | December | Teacher | 41-45 | Day-care $\leq 6$ years | 45 | Unknown | 10 | 5 | 5 |
| Cluster 4 | November | Teacher | 41-45 | Day-care $\leq 6$ years | 79 | 86% | 10 | 8 | 2 |
| Cluster 5 | October | Teacher | 21-25 | Day-care $\leq 6$ years | 87 | 85% | 10 | 3 | 7 |
| Cluster 6 | November | Teacher | 16-20 | Day-care $\leq 6$ years | 37 | Unknown | 10 | 3 | 7 |
| Cluster 7 | October | Teacher | 51-55 | Day-care $\leq 6$ years | 106 | 95% | 15 | 5 | 10 |
| Cluster 8 | October | Child | 11-15 | Secondary school | 166 | 100% | 7 | 7 | 0 |
| Cluster 9 | November | Teacher | 46-50 | Unknown | 56 | Unknown | 7 | 0 | 7 |

<sup>a</sup>Date of symptom onset for symptomatic cases, date of test for asymptomatic cases, <sup>b</sup>High risk defined as person who stayed face to face ( $<1.5$ meters) for 15 minutes or longer, or in the same room for 30 minutes or longer with a COVID-19-case, respectively.<sup>12</sup> In crowded or unclear situations, or when resources do not allow for an individual risk assessment, particularly in the context of schools and day-care centres, all members of a class or group may be classified as high risk contact persons.<sup>10</sup>

**Figure 3:**
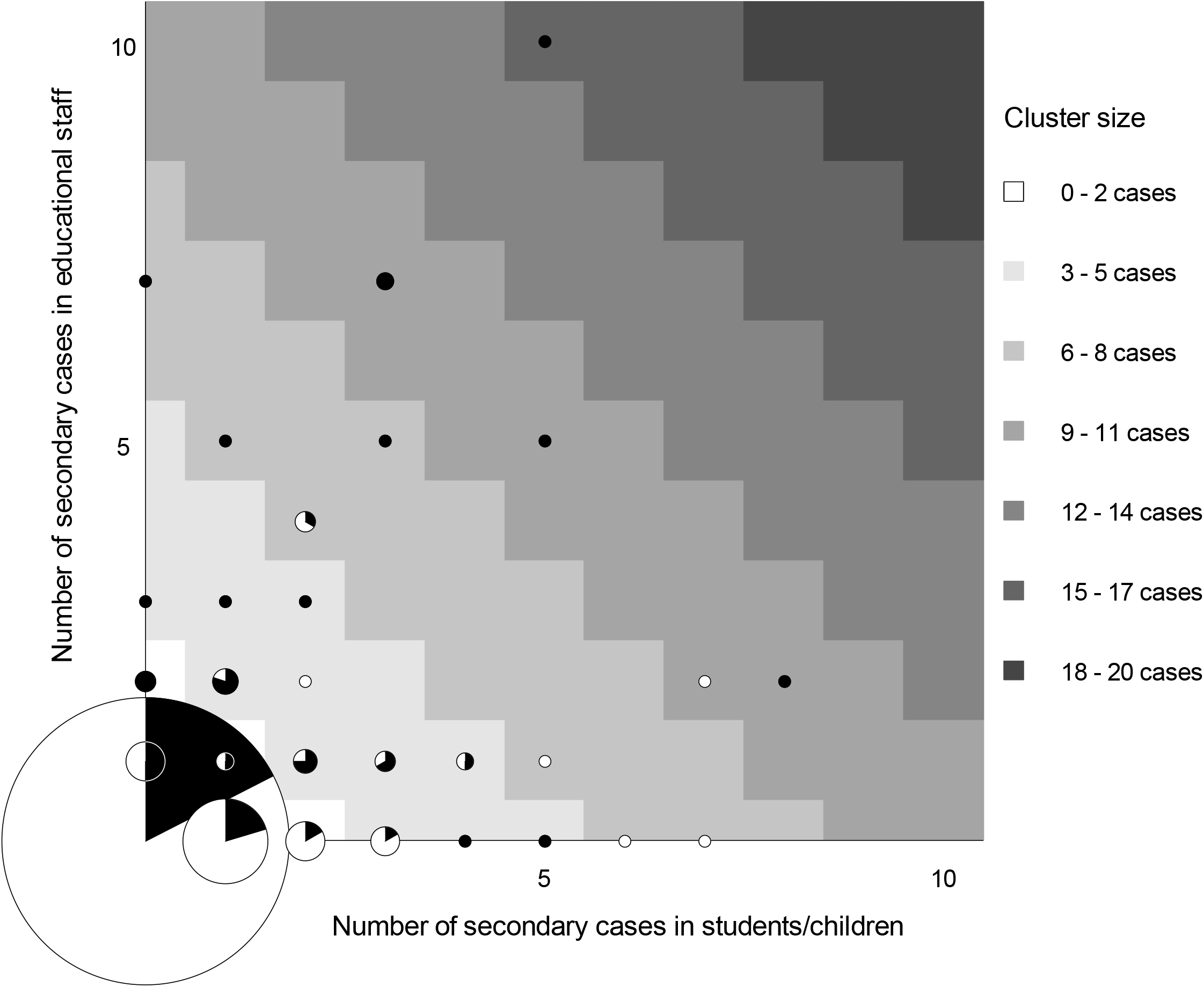
Frequency of secondary cases in children and teachers by role of the index case. Graph displays frequency and role of 784 index cases and their association with secondary transmission to teachers and students/children in schools and day-care centres in Rhineland-Palatinate, Germany, 2020. Grid position of circles represent the number of secondary cases in students/children (x-axis) and teachers (y-axis). The circle size is proportional to the number of index cases with the respective number of secondary cases reported in this study. The colour inside the circle represents the share of children (white) and teachers (black) observed among index cases represented by that circle. Circles in areas of the grid with same shade of grey represent clusters of similar size. For instance, the black dot at the very top of the grid identifies one cluster of size 15 (high cluster size = dark shade of grey) that emerged from a teacher index case (indicated by black colour vs. white colour of dot) and produced 10 secondary cases in teachers (position on y-axis) and 5 in student/children (position on x-axis).

## Discussion

This study provides evidence for an overall low SARS-CoV-2 transmission-risk in educational settings from August to December 2020 in Germany, with sharply increasing incidence rates, but with preventative measures in place. We found that approximately 1.3% of school contacts of an index case classified as category-I will become SARS-CoV-2 positive. When restricting the denominator to PCR-tested contacts, we found nearly the same secondary attack rate (SAR) of 1.5%. These numbers are well in line with other published findings on risk of transmission in schools.^16-21^ One other study from Germany based on 87 school index cases from the DPHA of Frankfurt calculated a slightly higher but comparable SAR of 1.9%.^16^ Other studies from different settings in Australia, Italy, Ireland, and Singapore also report comparable SARs between 0% and 3%.^17-20^

Compared to students/children, we found that the SAR was higher when the index case was a teacher. Likewise, the risk of causing a cluster and the mean number of secondary cases was higher when teachers were identified as index cases compared to students. Although not formally tested in other studies, mainly due to small sample size, descriptive findings in published literature already point towards larger numbers of secondary cases in teacher index cases and support our findings.^18,22,23^ In one study from the UK, half of eighteen primary school outbreaks involved teachers only.^22^ At the same time, we found that there is only limited transmission from child index cases to teachers, and that such events happen mainly in day-care centres: among a total of 591 student/child index cases, transmission was identified to 25 teachers, of which 22 were associated with thirteen children in day-care centres. The fact that teachers were the primary source of infection for teachers in day-care institutions supports the hypothesis that professional contacts independent of teaching activities with children (e.g. during staff meetings, work breaks) are likely to contribute to transmission.

The role of asymptomatic cases in the spread of the COVID-19 pandemic has been subject of an ongoing debate. A recent review and meta-analysis found that the proportion of asymptomatic cases was 17% of all COVID-19-positive cases.^24,25^ They further report that the risk of transmission was about 40% lower in asymptomatic cases as compared to symptomatic cases. While this is in line with our finding of a 50% lower attack rate in contacts of asymptomatic cases, we would like to interpret these with caution. Indeed, what we observed as ‘asymptomatic’ may in some cases just have been a PCR-diagnosis in the “pre-symptomatic” phase, e.g. in contact persons of COVID-19 positive cases outside the school/day-care setting. Hence, ‘asymptomatic’ cases in the presented study may in fact just have spent less days of their infectious period in school/day-care, thus explaining the lower risk of transmission. At the same time, these findings do not support the prevailing fear that asymptomatic cases could play a major role in the transmission of COVID-19 in schools/day-care under the current hygiene measures. One explanation for this finding, apart from shorter contact times due to ‘pre-symptomatic’ diagnosis, is a potential lower viral load in asymptomatic versus symptomatic cases.^26^

This study has limitations. First, we do not have a full survey of notified cases in the context of educational institutions from all sixteen reporting DPHAs. This raises the question of selection bias. However, we advised DPHAs to report consecutive index cases over at least a 4-week period or longer, thus reducing the chance of systematic under- or over-reporting of more or less salient index cases and associated under- or overestimation of transmission risk. Second, although all DPHAs routinely offer PCR tests to all contact persons to a COVID-19 index case at high-risk of transmission in the educational setting, 44% of our sample came from DPHAs that had outsourced sampling and testing to community testing centres. From these DPHAs, we received reliable information on secondary cases, but not on total contact persons and contact persons tested, since only positive test results are notifiable by testing-centres and associated laboratories. However, comparing the mean number of secondary cases between both samples shows similar results (0.39 versus 0.44 secondary cases per index case, data not shown), making us confident that these are from the same source population.

Third, the proportion of PCR tests among contact persons from DPHAs with internal organization of testing decreased in November and December, presumably as a function of increasing incidence and associated workload. However, only an estimated 20% of COVID-19 cases generally stay asymptomatic during the course of infection^24^ and all contact persons were monitored for disease symptoms. Thus, the 15% missing PCR tests in contact persons in November would result in 3% that could have become SARS-CoV-2 positive without being detected, making us confident that this had only minor effects on the presented results. Finally, our study attributes all transmissions in children/students and teachers detected in contact persons to COVID-19 index cases that were detected in educational settings to schools or day-care. This approach does not acknowledge that children/students and teachers may also have contact outside the institution, e.g. during leisure activities. This may have increased the presented risk estimates, particularly for child-to-child transmission, where exposure in the classroom and during leisure commonly coincides.

In summary, we could show that the risk of SARS-CoV-2 onwards transmission in the educational setting is low, but significantly higher, when teachers are index cases, mainly due to more common transmission between teachers in day-care centres. By contrast, there is lower risk of transmission between children/students, and only negligible risk for transmission from children/students to teachers in schools. We recommend a review of hygiene practices in the educational setting, with a focus on day-care centres and contact patterns between teachers. Besides, our findings also support the re-prioritization of vaccination to educational staff in day-care. These and similar measures guided by our findings have great potential to reduce the burden of infections, safe public health resources, and promote educational justice during the pandemic.

## Data Availability

Data and analytic code used for this study will be shared with researchers immediately following the publication in a peer-reviewed journal upon request from the corresponding author.

## Author contributions

The study idea, approach, and methods were conceptualised by PZ, MV, KJ, TB, and AS. PZ created the research questionnaire and the database. DH, CT, BV, SH, TK, KF, BK, SB, AM, KH, HM, AScha, and HK were responsible for the conduct of the study and for data collection. AS and PZ managed the database, verified the underlying data, performed statistical data analysis, and wrote the manuscript. AS provided the figures for the manuscript. All authors reviewed the manuscript, provided input, and approved the final version

## Data sharing

Data and analytic code used for this study will be shared with researchers immediately following the publication upon request from the corresponding author.

---

**Supplementary Figure 1:**
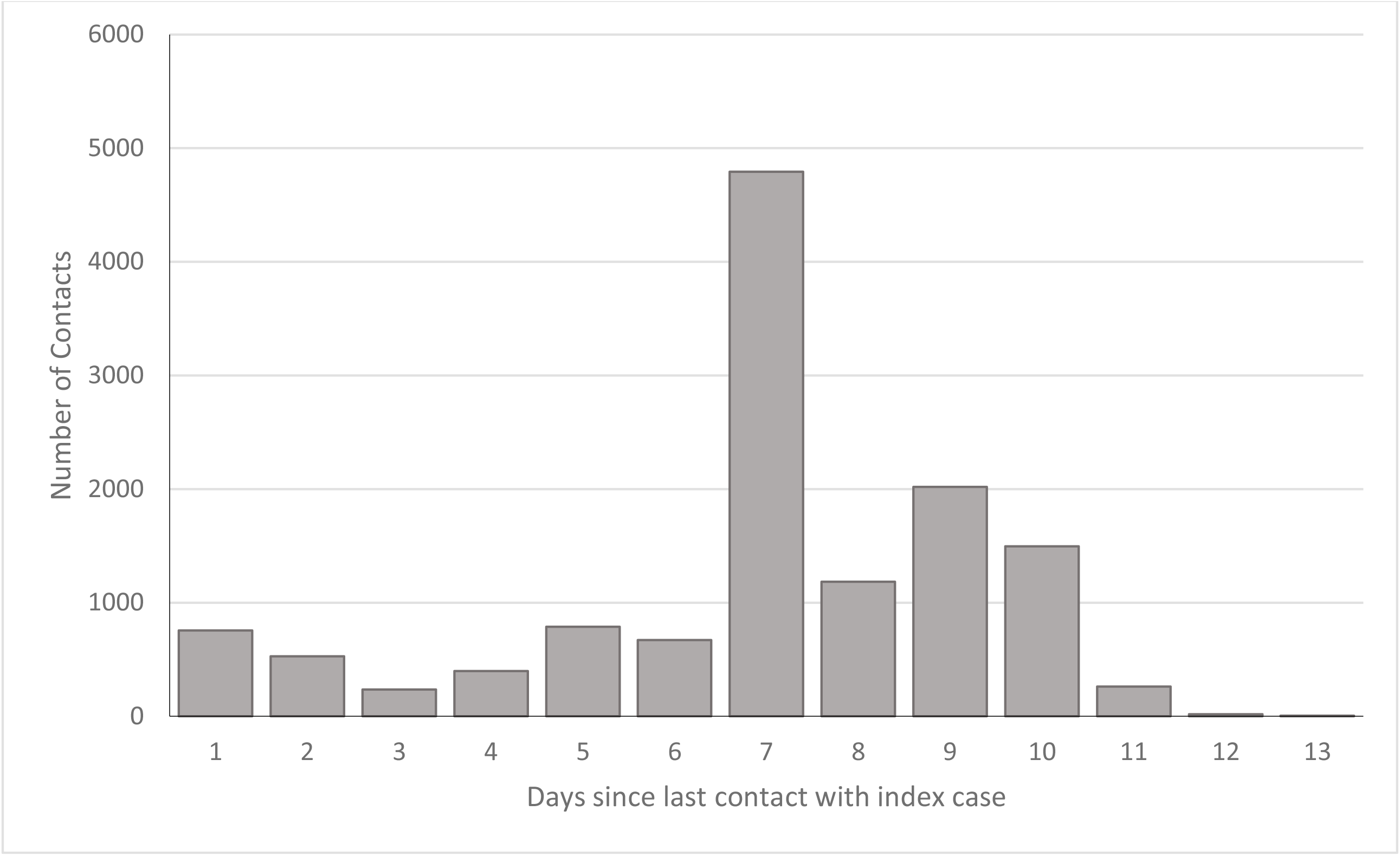
Timing of PCR testing in contact persons. Median time 7 days (Interquartile range: 6-9 days), 1.5% with missing information

**Supplementary figure 2:**
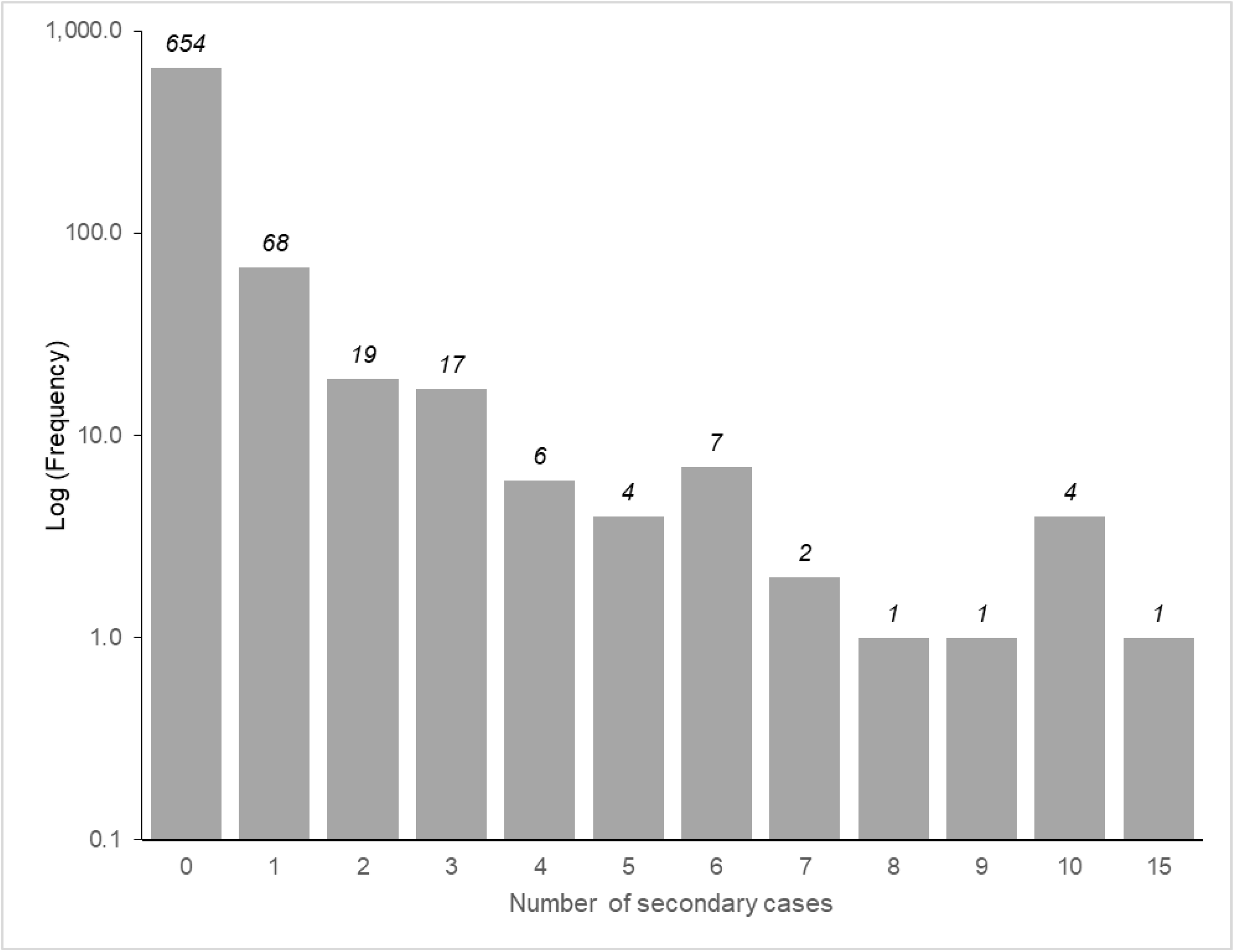
Cluster sizes. The median cluster size in those index cases who caused at least one secondary case was 1 (inter quartile range 1 to 3, minimum 1, maximum 15).

## STROBE Statement—checklist of items that should be included in reports of observational studies

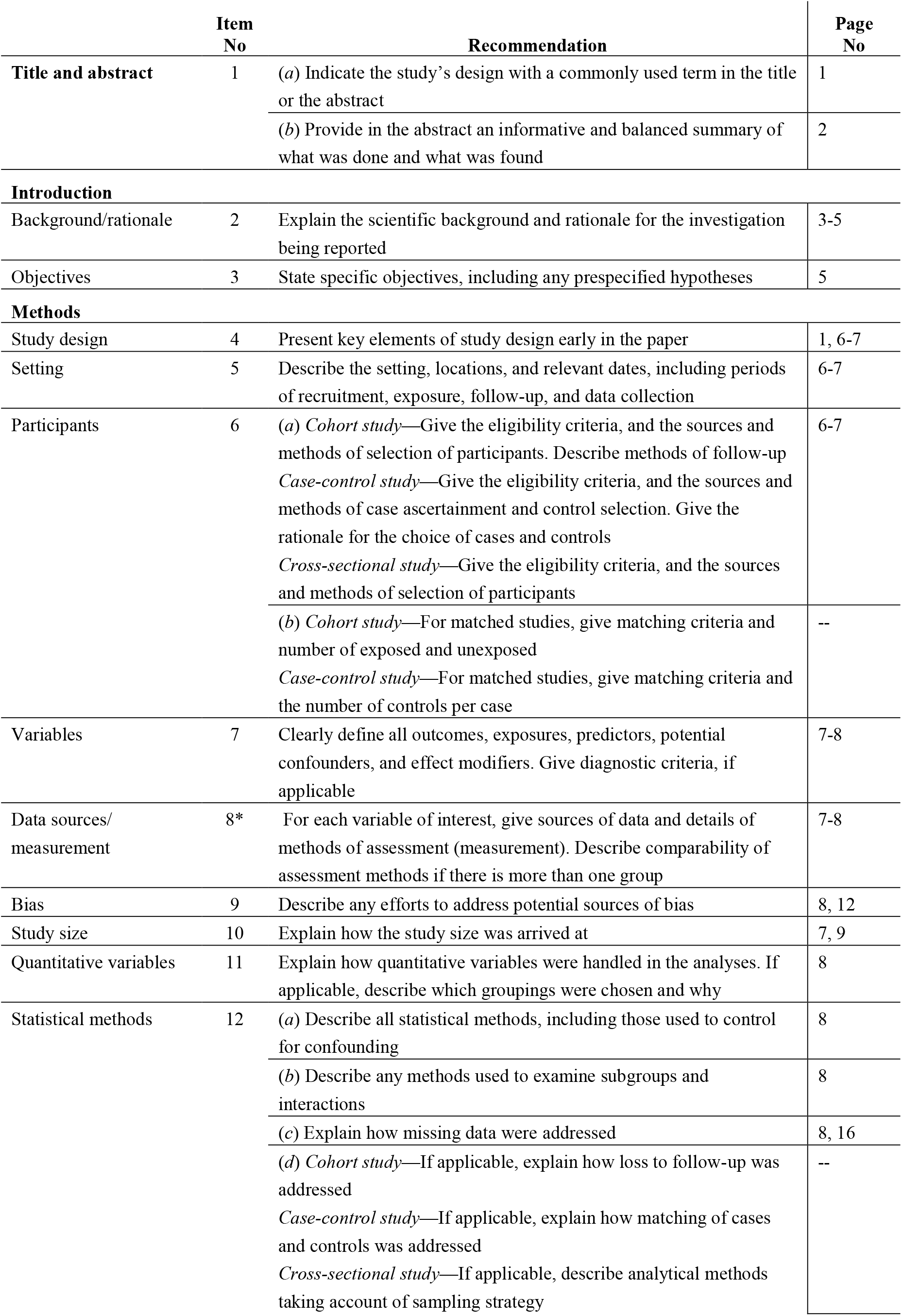
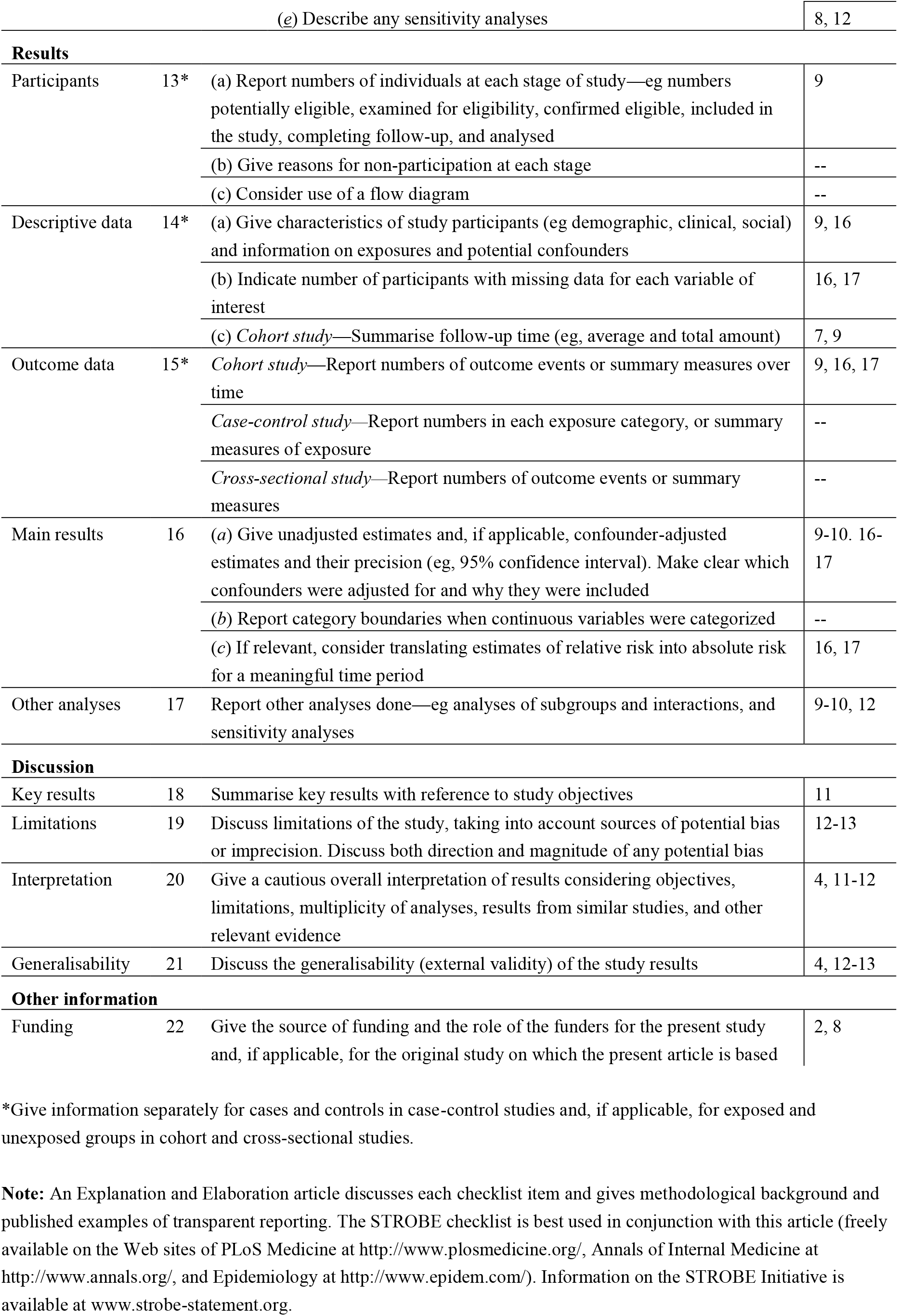

